# Uptake of COVID-19 vaccines among healthcare workers within primary healthcare facilities, Entebbe municipality Uganda

**DOI:** 10.1101/2022.10.20.22281300

**Authors:** N Kyakuwa, C Atuhairwe, H KalutTe, S Mpooya, F Nakanjako, L Perez, B Kikaire

## Abstract

**Background:** Routine vaccination is an essential highly successfully public health intervention in the prevention of infectious diseases that greatly depends on high coverage, and health care workers (HCWs) who play a pivotal role in ensuring the high uptake of vaccines in the population. COVID-19 vaccines have been proven efficacious, and vaccination campaigns have been ongoing, however, there is a perceived high vaccine hesitancy among health care workers in Uganda. This study describes the level and determinants of uptake of COVID-19 vaccines among HCWs in Entebbe municipality, Uganda.

**Materials and methods:** We conducted a health facility based cross-sectional study among HCWs from private and government health facilities in Entebbe municipality between July 2021 and August 2021. Structured questionnaires were used, and data were analysed using Stata version 12. We defined uptake as having received at least the first doze of COVID-19 vaccine or completed the two dozes.

**Results:** The level of vaccine uptake was 65.6%with higher uptake among males than females. HCWs aged 30-39 years were 2.7 times more likely to have been vaccinated than those less than 30 years (OR 2.72, 95% CI: 1.26-5.88, P-value <0.01), and the odds of having been vaccinated were 4 times higher among health workers above 40 years (OR 4.29, 95% CI 1.50-12.24, P-value < 0.01). Additionally, the odds of having been vaccinated were 4 times higher among health care workers that participated in COVID-19 vaccine related activities (OR 4.18, 95% CI 2.16-8.10, p-value <0.001). Healthcare workers (98%) had confidence in the vaccines although 45% of those that were not vaccinated felt that the vaccines were ineffective.

**Conclusion:** Vaccine uptake among HCWs was relatively high compared to the WHO recommended uptake of 70% by mid-2022, although some HCWs were still hesitant. The convenience of vaccination services was an important factor in vaccine uptake. Hence, governments should endeavour to improve access to vaccination both for HCWs and the public.

## Background

Vaccination is one of the cost-effective public health interventions in the control of infectious diseases in populations (1). The incidences of childhood vaccine preventable diseases such as measles, polio, pertussis have drastically decreased due to global vaccination campaigns (1, 2), and other infectious diseases such as influenza have been reduced through periodic vaccination programs (3, 4). Besides the direct effect of protection to the vaccinated individual, high vaccination coverage rates provide indirect benefits to the community through herd immunity (5, 6). This protection is important in reducing transmission rates thus decreasing the risk of infections among susceptible, un-vaccinated individuals within the community (7).

Although vaccination is globally accepted as one of the most successful measures in the control of infectious diseases, (8, 9) vaccine hesitancy persists in the population and health care workers(HCWs) specifically (10) with some individuals perceiving vaccines as unsafe and unnecessary (11). Highest scepticism has been reported among those with highest level of education (12). Vaccine hesitancy among HCWs remains a public health threat (13), and is highly context-, vaccine-, and profession specific (14). Unvaccinated HCWs are at a risk of contracting infections from their patients, and similarly, patients could contract infections from HCWs. A study carried out among HCWs in Los Angeles showed that acceptance of COVID-19 vaccine varied with the role of HCWs with physicians and research scientists being more likely to take vaccines than others (15).

Globally, vaccination against SARS-COV-2 met hesitancy or low uptake which threatens the attainment of the WHO recommended uptake of 70% by mid-2022. Additionally, the relatively high risk of severe corona virus disease among the elderly and those with comorbidities increases COVID-19 vaccine hesitancy among those perceived to be at low risk of severe disease. COVID-19 vaccine acceptance rates vary from region to region with Africa being one of the regions found with low rates of COVID-19 vaccine acceptance (16). A worldwide systematic review about COVID-19 vaccine acceptance among HCWs showed vaccine acceptance rates ranging from 27.7% in the Democratic Republic of the Congo to 78.1% in Israel (16) with many countries falling in between. The highest vaccine hesitancy to COVID-19 was reported in Cameroon (17). The major reasons reported by the hesitant people included; (i) being against vaccines in general; (ii) concerns about safety (thinking that a vaccine produced in a rush is too dangerous); (iii) considering the vaccine useless because of the harmless nature of COVID-19; (iv) general lack of trust; (v) doubts about the efficiency of the vaccine; (vi) belief to be already immunized; and (vii) doubt about the provenance of vaccine (18). While several studies have explored intentions to be vaccinated in the general population, few studies have assessed actual vaccine uptake, more so among HCWs. Therefore, the study assessed the level of uptake of COVID-19 vaccines among HCWs within Entebbe municipality in Uganda.

## Materials and methods

### Study Design, Setting and Participants

A cross-section study design was used to assess the level of uptake and determinants of COVID-19 vaccine uptake among HCWs from Entebbe Municipality Wakiso District in Uganda between July and August 2021. Entebbe town has a population of 67 271 people. The municipality has 1 research centre, and 40 healthcare facilities 33 of which are privately owned while 7 are government owned facilities. The government health facilities are distributed as follows; i) one regional referral hospital, ii) one health centre IV, iii) three health centre III, and iv) 2 health centre II. The study was carried out in both private and primary health care government health facilities (II, III, & IV) including health research centers within Entebbe Municipality. The regional referral hospital was excluded due to a similar study that was ongoing at that time.

COVID-19 vaccination schedules in Entebbe was first prioritized for the elderly above 45 years, those with other chronic illness, and workers in risky environments including healthcare workers. But later extended to all above 18years of age.

Participants were mainly HCWs providing direct clinical care to patients in community, health facility and or research centers. These included medical doctors, nurses, nursing assistants, allied health professionals, social workers, research scientists and other roles involving direct patient interaction. The study also included all non HCWs not involved in providing clinical services in health facilities and research centers, such as managers, receptionists, other administrative roles, cleaners, porters, janitors and other non-clinical roles.

Sample size was determined to be 304 participants using Daniel 1999 formula for one sample, using vaccine hesitance rate of 72,3% based on a study by Sellam et al(16) in Democratic Republic of Congo (DRC). The study was conducted during lockdown period and healthcare workers were working in shifts. Therefore, the anticipated non respondence was 20% (61participants) leading to a total 364 participants.

### Data source

Data was collected using a structured questionnaire that was distributed either as a hard copy or electronically and stored and managed according to Uganda Virus Research Institute (UVRI) data management guidelines.

Level of uptake of COVID-19 vaccines for the different HCWs was reported as proportions, and the determinants of COVID-19 vaccine uptake were analyzed using logistic regression.

The study was conducted according to ICH/GCP and the national and international regulations for research in humans.

Ethical approval was obtained from UVRI Research Ethics Committee (REC).

Informed consent was sought from all participants. All information was kept confidential and safe under lock and key.

### COVID-19 risk management

Research Assistants were provided with face masks and handed sanitizers throughout the study. Both the research assistant and the participants wore masks during interviews. During interviews a physical distance of at least two meters was maintained between the participants and the research assistant. As much as possible interviews were conducted in an open space.

### Measurements

The study outcome was uptake of COVID-19 vaccination either as initiation or completion.

The independent variables were the social demographics, healthcare level of service, profession, level of education, type of facility, testing for COVID-19, participation in COVID-19 vaccination activities, and having cared for a COVID-19 patient.

### Statistical analysis

Data was analyzed using stata version 12

## Results

### Level of uptake of COVID-19 vaccines

Overall, 360(98.5%) healthcare workers in Entebbe municipality participated in study 236 (65.6%) of whom had been vaccinated.

### Socio-demographic characteristics

More than half of the participants were female 222 (61.7%) With a mean age of 31.0 years (SD± 7.95). Most of the participants had a Bachelors/diploma 263 (73.1%), 66 (18.3%) had completed Secondary level and 24(6.7%) had a Masters’ degree. Majority 248 (68.9%) of the study participants were medical workers such as medical officers, nurses, dentists among others, while the non-medical workers included accountants, administrators, and security personnel. Of the 248 medical workers; 39.9% were nurses, 21.0% were laboratory personnel, 10.9% were clinical officers (diploma level clinician), 8.8% were medical doctors and 19.3% belonged to other medical fields that included radiologists, pharmacists, physiotherapists, dental officers, and nutritionists. On the other hand, among 112 non-medical workers, more than half (52.6%) were support staff whereas 47.4% had administrative roles. Table 1 shows the social demographic factors of the respondents.

**Table 1.** Socio-demographic characteristics of respondents (N=360)

| Category | Vaccine uptake |  |  |
| --- | --- | --- | --- |
|  | Total (%) | No (%) | Yes (%) |
| **Gender** |  |  |  |
| Female | 222 (61.7) | 71 (57.3) | 151 (64.0) |
| Male | 138 (38.3) | 53 (42.7) | 85 (36.0) |
| **Adult Age group** |  |  |  |
| 18-29 Years | 191 (53.1) | 83 (66.9) | 108 (45.8) |
| 30-39 Years | 111 (30.8) | 31 (25.0) | 80 (33.9) |
| >40 years | 58 (16.1) | 10 (8.1) | 48 (20.3) |
| **Level of education** |  |  |  |
| Primary | 7 (1.9) | 4 (3.2) | 3 (1.3) |
| Secondary | 66 (18.3) | 25 (20.2) | 41 (17.4) |
| Diploma/Bachelors | 263 (73.1) | 92 (74.2) | 171 (72.5) |
| Masters | 24 (6.7) | 3 (2.4) | 21 (8.9) |
| **Job category** |  |  |  |
| Medical | 248 (68.9) | 81 (65.3) | 167 (70.8) |
| Non-medical | 112 (31.1) | 43 (34.7) | 69 (29.2) |
| **Cadre/job title(n=248)** |  |  |  |
| Consultant/senior consultant/Professor | 3 (1.2) | 1 (1.2) | 2 (1.2) |
| Medical officer special grade/registrar | 6 (2.4) | 0 (0.0) | 6 (3.6) |
| Medical officer/Dental surgeon | 12 (4.8) | 3 (3.7) | 9 (5.4) |
| Intern doctor | 1 (0.4) | 0 (0.0) | 1 (0.6) |
| Clinical officer/Paramedic | 27 (10.9) | 8 (9.9) | 19 (11.4) |
| Nursing officer/midwife | 99 (39.9) | 30 (37.0) | 69 (41.3) |
| Laboratory technologist | 52 (21.0) | 25 (30.9) | 27 (16.2) |
| Dental officer | 3 (1.2) | 1 (1.2) | 2 (1.2) |
| Radiologist, Physiotherapists, pharmacists & nutritionists | 45 (18.1) | 13 (16.0) | 32 (19.2) |
| **Non-medical (n=112)** |  |  |  |
| Receptionist | 13 (11.6) | 6 (14.0) | 7 (10.1) |
| Cashier | 6 (5.4) | 2 (4.7) | 4 (5.8) |
| Administrator | 29 (25.9) | 10 (23.3) | 19 (27.5) |
| Accountant | 5 (4.5) | 2 (4.7) | 3 (4.3) |
| Cleaner | 19 (17.0) | 6 (14.0) | 13 (18.8) |
| Security | 14 (12.5) | 8 (18.6) | 6 (8.7) |
| Driver | 4 (3.6) | 1 (2.3) | 3 (4.3) |
| Storekeeper/procurement | 2 (1.8) | 2 (4.7) | 0 (0.0) |
| Executives & gardeners | 20 (17.9) | 6 (14.0) | 14 (20.3) |

The distribution of health workers’ areas of operation is shown in Table 2. One hundred and three (28%) of the respondents worked in the out-patients department. Other work-areas included maternity 53 (14.7%), in-patient ward 19 (5.2%), operating theatres 1 (0.3%), intensive care unit 2 (0.6%), laboratory 59 (16.4%), ART clinic 12 (3.3%), non-clinical area 9 (2.5%), administrative offices 47 (13.0%) and isolation rooms 1 (0.03%). Regarding the level of service, the health workers worked at the following establishments: hospital 54 (15.0%), Health Centre IV 3 (0.8%), Health Centre II & III 144 (37.9%), medical centre 61 (16.9%), and private clinics 98 (27.2%).

**Table 2.** Distribution of health service-related factors (N=360)

| Category | Vaccine uptake |  |  |
| --- | --- | --- | --- |
|  | Total (%) | No (%) | Yes (%) |
| **Area of operation** |  |  |  |
| Out-patient | 103 (28.6) | 34 (27.4) | 69 (29.2) |
| Maternity/Antenatal | 53 (14.7) | 18 (14.5) | 35 (14.8) |
| In-patient ward | 19 (5.3) | 1 (0.8) | 18 (7.6) |
| Operating theatre | 1 (0.3) | 0 (0.0) | 1 (0.4) |
| Intensive Care Unit | 2 (0.6) | 0 (0.0) | 2 (0.8) |
| Laboratory | 59 (16.4) | 28 (22.6) | 31 (13.1) |
| ART clinic | 12 (3.3) | 3 (2.4) | 9 (3.8) |
| Non-clinical area | 9 (2.5) | 4 (3.2) | 5 (2.1) |
| Administrative office | 47 (13.1) | 14 (11.3) | 33 (14.0) |
| Isolation wards/rooms | 1 (0.3) | 0 (0.0) | 1 (0.4) |
| Others | 54 (15.0) | 22 (17.7) | 32 (13.6) |
| **Service providers** |  |  |  |
| Hospital | 54 (15.0) | 17 (13.7) | 37 (15.7) |
| Health centre IV | 3 (0.8) | 0 (0.0) | 3 (1.3) |
| Health centre II & III | 144 (37.9) | 30 (20.8) | 114 (79.2) |
| Medical centre | 61 (16.9) | 26 (21.0) | 35 (14.8) |
| Private clinic | 98 (27.2) | 51 (41.1) | 47 (19.9) |
| **Type of hospital ownership** |  |  |  |
| Private not for profit (PNFP) | 84 (23.3) | 39 (31.5) | 45 (19.1) |
| Private for profit (PFP) | 146 (40.6) | 72 (58.1) | 74 (31.4) |
| Government | 130 (36.1) | 13 (10.5) | 117 (49.6) |
| **Had ever cared for COVID-19 patients** |  |  |  |
| No | 134 (37.2) | 54 (43.5) | 80 (33.9) |
| Yes | 226 (62.8) | 70 (56.5) | 156 (66.1) |
| **Had ever tested for COVID-19** |  |  |  |
| No | 89 (24.7) | 58 (46.8) | 31 (13.1) |
| Yes | 271 (75.3) | 66 (53.2) | 205 (86.9) |
| **If YES, what were the results?** |  |  |  |
| Negative | 230 (84.9) | 53 (80.3) | 177 (86.3) |
| Positive | 41 (15.1) | 13 (19.7) | 28 (13.7) |
| **Took part in COVID-19 vaccine activities** |  |  |  |
| No | 150 (41.7) | 91 (73.4) | 59 (25.0) |
| Yes | 210 (58.3) | 33 (26.6) | 177 (75.0) |

One hundred and forty-six (40%) of the health workers worked for Private-For-Profit (PFPs) health facilities, while one hundred and thirty (36%) worked at government-run health facilities.

### Determinants of COVID-19 vaccine uptake among healthcare workers in Entebbe

#### Socio-demographic characteristics

At bivariate analysis, the odds of being vaccinated were almost 10 times higher among health workers who were 40 years and more (OR 9.33, 95% CI 1.36-63.60, P value = 0.02). The odds of being vaccinated were 0.9 times lower among health workers who were female, but this was not statistically significant (OR 0.93, CI 0.57-1.53, P value = 0.78). No other socio-demographic factor was found to have a signification association with vaccine uptake (Table 3).

**Table 3.** Bivariate analysis of socio-demographic characteristics

| Category | Vaccine Uptake |  |  | OR (95% CI) | p-value |
| --- | --- | --- | --- | --- | --- |
|  | No (%) | Yes (%) | Total (%) |  |  |
| **Gender** |  |  |  |  |  |
| Male | 53 (38.4) | 85 (61.6) | 138 (100.0) | Ref. |  |
| Female | 71 (32.0) | 151 (68.0) | 222 (100.0) | 0.93 (0.57-1.53) | 0.78 |
| **Adult Age group** |  |  |  |  |  |
| 18-29 Years | 83 (43.5) | 108 (56.5) | 191 (100.0) | Ref. |  |
| 30-39 Years | 31 (27.9) | 80 (72.1) | 111 (100.0) | 2.48 (0.54-11.31) | 0.24 |
| >40 years | 10 (17.2) | 48 (82.8) | 58 (100.0) | 9.33 (1.36-63.96) | 0.02 |
| **Level of education** |  |  |  |  |  |
| Primary | 4 (57.1) | 3 (42.9) | 7 (100.0) | 0.55 (0.10-3.12) | 0.50 |
| Secondary | 25 (37.9) | 41 (62.1) | 66 (100.0) | 0.68 (0.25-1.86) | 0.45 |
| Diploma/Bachelors | 92 (35.0) | 171 (65.0) | 263 (100.0) | 1.59 (0.63-4.04) | 0.33 |
| Masters | 3 (12.5) | 21 (87.5) | 24 (100.0) | Ref. |  |
| **Job category** |  |  |  |  |  |
| Medical | 81 (32.7) | 167 (67.3) | 248 (100.0) | 1.53 (0.93-2.53) | 0.10 |
| Non-medical | 43 (38.4) | 69 (61.2) | 112 (100.0) | Ref. |  |
| **Cadre/job title(n=248)** |  |  |  |  |  |
| Consultant/senior consultant/Professor | 1 (33.3) | 2 (66.7) | 3 (100.0) | 0.43 (0.04-5.37) | 0.51 |
| Medical officer special grade/registrar | 0 (0.0) | 6 (100.0) | 6 (100.0) | 1.08 (0.11-10.56) | 0.95 |
| Medical officer/Dental surgeon | 3 (25.0) | 9 (75.0) | 12 (100.0) | 1.08 (0.20-5.92) | 0.93 |
| Intern doctor | 0 (0.0) | 1(100.0) | 1 (100.0) | - | - |
| Clinical officer/Paramedic | 8 (29.6) | 19 (70.4) | 27 (100.0) | 0.75 (.231-2.478) | .645 |
| Nursing officer/midwife | 30 (30.3) | 69 (69.7) | 99 (100.0) | .714 (.292-1.749) | .46 |
| Laboratory technologist | 25 (48.1) | 27 (51.9) | 52 (100.0) | .649 (.241-1.744) | .391 |
| Dental officer | 1 (33.3) | 2 (66.7) | 3 (100.0) | .432 (.035-5.370) | .514 |
| Radiologist, pharmacists & nutritionists | 13 (28.9) | 32 (71.1) | 45 (100.0) | Ref. |  |
| **Non-medical (n=112)** |  |  |  |  |  |
| Receptionist | 6 (46.2) | 7 (53.8) | 13 (100.0) | .292 (.062-1.368) | .118 |
| Cashier | 2 (33.3) | 4 (66.7) | 6 (100.0) | .50 (.066-3.770) | .501 |
| Administrator | 10 (34.5) | 19 (65.5) | 29 (100.0) | 1.562 (.341-7.154) | .565 |
| Accountant | 2 (40.0) | 3 (60.0) | 5 (100.0) | - | - |
| Cleaner | 6 (31.6) | 13 (68.4) | 19 (100.0) | .278 (.067-1.147) | .077 |
| Security | 8 (57.1) | 6 (42.9) | 14 (100.0) | .250 (.055-1.138) | .073 |
| Driver | 1 (25.0) | 3 (75.0) | 4 (100.0) | .750 (.061-9.270) | .823 |
| Storekeeper/procurement | 2 (100.0) | 0 (0.0) | 2 (100.0) | - |  |
| Gardeners, executives | 6 (30.0) | 14 (70.0) | 20 (100.0) | Ref |  |

†95% confidence intervals for odds ratios (OR) are in brackets, ref. reference category
\*p<0.05 at 5% level of significance, (-) OR could not be computed

#### Health service-related factors

Among the health service-related factors, working in the in-patient wards increased the odds of having been vaccinated by more than 120 times (OR 12.38, 95% CI 1.54-99.61), P-value 0.018. However, the 95% CI is wide, and these should be interpreted with caution. Similarly, working in a hospital (OR 2.36, 95% CI 1.18-4.75), P-value 0.016 and working at a health II or III level (OR 4.12, 95% CI 2.34-7.25), P-value <0.01 increased the odds of having been vaccinated.

The health workers in privately-run health care facilities (Private-For-Profit & Private-Not-For-Profit) were less likely to have been vaccinated compared to government facilities. The odds of having been vaccinated were 87% less in the private not for profit (OR 0.13, 95% CI 0.06 – 0.33) P-value <0.001, and almost 90% less in private for-profit facilities (OR 0.09, 95% CI 0.03-0.20), P-value <0.001. More associations of health service-related factors and uptake of vaccines are in Table 4.

**Table 4.**
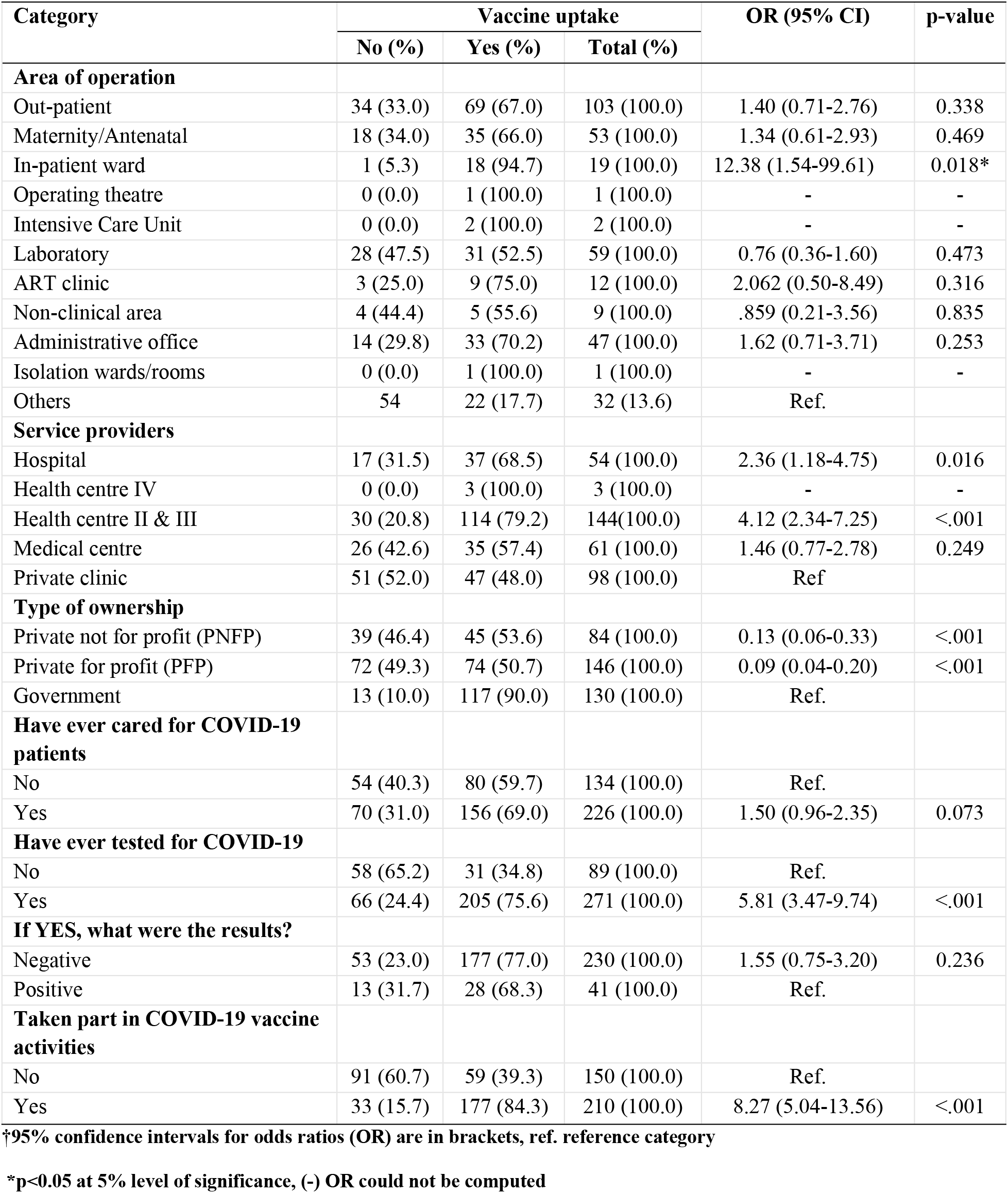
Health service factors and association with uptake of COVID-19 vaccines.

At Multivariate analysis, respondents that were aged 30-39 years were 2.7 times more likely to have been vaccinated than those less than 30 years (OR 2.72, 95% CI: 1.26-5.88, P value <0.01), and the odds of having been vaccinated were 4 times higher among health workers above 40 years (OR 4.29, 95% CI 1.50-12.24, P value < 0.01). Additionally, the odds of having been vaccinated were 4 times higher among health care workers that participated in COVID-19 vaccine related activities (OR 4.18, 95% CI 2.16-8.10, p-value <0.001).

The HCWs that worked in Private-Not-Profit (PNFP) organizations were less likely to have been vaccinated (OR 0.23; 95% CI: 0.07-0.53, P value <0.001) or for Private-For-Profit (PFP) organizations (OR 0.19; 95% CI: 0.05-0.48, P value <0.001).

A health worker with a negative test result was 1.7 times more likely to have been vaccinated than one with a positive result (OR 1.79, 95% CI: 0.74-4.32, P value 0.12), however, this was not significant. Table 5 has the details of the multivariate analysis.

**Table 5.** Determinants of COVID-19 uptake.

| Categories | Odds Ratio | 95% C.I. for Odds Ratio | P-value |
| --- | --- | --- | --- |
| **Gender (male)** | 1.985 | 1.006-3.916 | 0.048 |
| **Age group** |  |  |  |
| 18-29 | 1 |  |  |
| 30-39 Years | 2.72 | 1.26-5.89 | 0.011 |
| +40 Years | 4.29 | 1.50-12.24 | 0.006 |
| **Type of ownership** |  |  |  |
| Government | 1 |  |  |
| Private not for profit (PNFP) | 0.23 | 0.09-0.57 | <0.001 |
| Private for profit (PFP) | 0.19 | 0.08-0.42 | <0.001 |
| **Had cared for a coronavirus patient** |  |  |  |
| Yes | 0.73 | 0.37-1.45 | 0.365 |
| **COVID-19 test results** |  |  |  |
| Negative | 1.79 | 0.74-4.32 | 0.199 |
| **Participated in any COVID-19 vaccine related activities** |  |  |  |
| Yes | 4.18 | 2.16-8.11 | <0.001 |

95% confidence intervals for odds ratios (OR), \*p<0.05 at 5% level of significance

## Discussion

In this study, the uptake of COVID-19 vaccines among HCWs within Entebbe municipality was 65.6%. This uptake is close to the WHO globally recommended 70% vaccine uptake level by mid-2022 and the finding of Lubega et al 2021 who reported an uptake of up to 70% among health care workers in two hospitals in Uganda (19). Whereas Lubega studied health care workers in tertiary hospitals, this study was conducted among health care workers in primary health care units thus the differences in uptake of the vaccines. Our findings differ from a study in Nigeria that reported uptake rate of 33% that completed two doses of the COVID-19 vaccine (20). The Nigeria study was conducted in the early days of COVID-19 vaccine introduction, when many unknowns about the vaccine existed, while our study was carried out much later and after the highly fatal increase in mortality due to the delta variant of the COVID-19 pandemic. It is probable that the relatively high uptake of vaccines by HCWs observed in our study was driven by the fear resulting from the ‘delta wave’. A multi-ethnic study in the UK healthcare workforce by Christopher A. Martins et al 2021 reported uptake rates of 64.5% (21), and this is similar to the findings of our study. A study by Maria L. Pacella-LaBarbara et al 2021 among emergency HCWs in US (22) reported vaccine uptake of 79% which is much higher than what we found. This could be due to differences in risk perceptions by the HCWs in the two studies. Unlike in our study where the mean age was 31 years, the mean age study of participants in Maria’s was 41 years. It is well known that the risk of severe disease increases with increased age, therefore the difference in age could have led to a higher uptake in Maria’s study compared to ours. Furthermore, 29% of the participants had underlying health conditions, a variable that was not assessed for in this study.

Most studies both in Africa and globally have explored vaccine acceptance among HCWs and reported acceptance rates ranging between 50-70% (23-26). In Pakistan, vaccine acceptance was reported at 70.2% (27) while in Canada, Stefania Dzieciolowska et al 2021 reported vaccine acceptance rate of 80.9% (28), and China reported vaccine acceptance of 86.2% (29). While these studies showed that health care workers were likely to take up COVID-19 vaccines once available, it should be noted that the actual uptake may be different from the intention (30, 31). Studies in the United Kingdom of Saudi Arabia showed that the intention of HCWs taking vaccination was 70%, but when they studied actual uptake, the acceptance rate was 33.2% (26, 32). However as noted above few studies have explored the actual vaccine uptake and this requires further investigation.

We found that being at least forty years increased the odds of taking up the COVID-19 vaccines. This is not surprising since the disease in known to preferentially affect the older people and other studies have reported similar findings (33-36). This age related perception of risk also explains why younger health care workers were less likely to be vaccinated, since the risk of severe disease has been reported to be low among younger age groups (37) although some studies have reported severe diseases among younger age groups. Andrea et al reported that almost 70% of the young ‘low risk’ with ongoing symptoms of COVID-19 had impairment in one or more organs four months after initial symptoms of SARS-CoV-2 infection (38). This study seems to suggest that even though low risk individuals have lower mortality rates, their severity of disease is seen by organ temporary impairment and follow up of this category is of public health importance.

The findings of our study showed that the men were more likely to be vaccinated as compared to women, which agrees with other studies (22, 25). Just like the age related risk perception, the male gender has been reported to suffer more severe acute respiratory distress syndrome compared to females (39-41) and this could have driven more men to seek vaccination. Although a study in Western Uganda reported that having attained tertiary level of education increased the intention to be vaccinated (25) and Megan Halbrook et al, 2021 in California reported that vaccine uptake was associated with higher level of education (42), we found that level of education did not affect uptake of vaccines among health care workers in Entebbe. Level of education may increase perception of risk, and since HCWs have a good understanding of risk, their education level may thus not be a factor affecting actual uptake of vaccination.

Working at the in-patient departments was associated with increased uptake of the vaccines. Being hospitalized is an indicator of disease severity, hence, HCWs working in these departments care for more sick patients than their counterparts at out-patients. This brings about differences in risk perceptions. Having cared for patients suffering from COVID-19 also increased the likelihood of being vaccinated. This further emphasizes the risk perception.

The convenience/reliability of vaccination services is critical in vaccine uptake. In this study, we found out that being a government worker was associated with increased uptake of vaccines than working in private for profit and private not for profit healthy facilities. Rollout of COVID-19 vaccination in Uganda has been majorly in government hospital. This accessibility to vaccination services could have led to higher uptake among HCWs in these facilities. Similar studies have reported that accessibility to vaccine services played a key role in completion of vaccination schedules in children (43). The participation in COVID-19 vaccine related activities was associated with increased uptake of the vaccines. Participating in such activities builds vaccine confidence. HCWs who have been vaccinated are more likely to encourage their clients/patients to get vaccinated than the hesitant workers. These workers are trained and given more information about the vaccines which further builds confidence and trust in vaccines. However, no studies have been done in this area hence a need for further research.

The study further explored the relationship between previous testing for SARS-COV-2 and vaccine uptake among HCWs. We found out that HCWs who had ever tested for SARS-COV-2 virus were more likely to take the vaccines than their counter parts. Similar findings were reported by Mazin Barry et al 2021 Participants who tested negative were more likely to take the vaccine. Testing positive was possibly taken as natural immunization, therefore, the HCWs didn’t see the need to be vaccinated. More to this, there has been theoretical belief of immune enhancement of disease implying that those that tested positive could have feared to take the vaccine.

The study was carried out during lock down, and this was a limitation since participants were mainly reached via internet. Their assertion of having been vaccinated couldn’t be verified. Random sampling was not possible due to prioritization of only essential workers and also working in shifts.

## Conclusion

The study reported a high level of uptake of vaccines among HCWs within Entebbe municipality. However, there remains a significant proportion of HCWs who are hesitant to take vaccination and further studies are needed to understand and better address the reasons for the vaccine hesitancy.

The study found a relatively moderate uptake of vaccines among HCWs in Entebbe municipality. Confidence in vaccines and an enabling work environment are among the reasons that increased vaccine uptake. However, there remains a significant proportion of HCWs that is hesitant to take the COVID-19 vaccines and HCWs combined efforts to reduce this proportions are needed.

## Recommendation

This study found out that uptake was improved by vaccine accessibility and participation in COVID-19 vaccination services. Hence, the government should ensure that vaccination services are accessible to HCWs and the general public. The involvement of private health facilities in the COVID-19 vaccination campaign would improve vaccine uptake both among HCWs and the general public. Further longitudinal studies involving multi stage health professionals exploring determinants of vaccine uptake is recommended as this will elicit an understanding of the drivers of vaccine hesitancy among HCWs.

## Data Availability

Data cannot be shared publicly because of Country specific data sharing restrictions. Data are available from the UVRI Institutional Data Access / Ethics Committee (contact via +256773747607) for researchers who meet the criteria for access to confidential data.

## Acknowledgment

We thank our study participants for accepting to take part in this study.

